# Humoral and cellular immune correlates of protection against COVID-19 in kidney transplant recipients

**DOI:** 10.1101/2022.08.21.22279029

**Authors:** D. Kemlin, N. Gemander, S. Depickère, V. Olislagers, D. Georges, A. Waegemans, P. Pannus, A. Lemy, M. E. Goossens, I. Desombere, J. Michiels, M. Vandevenne, L. Heyndrickx, K.K. Ariën, A. Matagne, M.E. Ackerman, A. Le Moine, A. Marchant

## Abstract

As solid organ recipients are at high risk of severe COVID-19 and respond poorly to primary SARS-CoV-2 mRNA vaccination, they have been prioritized for booster vaccination. However, an immunological correlate of protection has not been identified in this vulnerable population. We conducted a prospective monocentric cohort study of 65 kidney transplant recipients who received three doses of SARS-CoV-2 BNT162b2 mRNA vaccination. Associations between symptomatic breakthrough infection (BTI) and vaccine responses, patient demographic and clinical characteristics were explored. Symptomatic COVID-19 was diagnosed in 32% of kidney transplant recipients during a period of six months after the administration of the third vaccine dose. During this period, SARS-CoV-2 delta and omicron were the dominant variants in the general population. Univariate analyzes identified avidity of SARS-CoV-2 receptor binding domain (RBD) binding IgG, neutralizing antibodies and SARS-CoV-2 S2 domain-specific IFN-γ responses as correlates of protection against BTI. Some demographic and clinical parameters correlated with vaccine responses, but none correlated with the risk of BTI. In multivariate analysis, the risk of BTI was best predicted by neutralizing antibody and S2-specific IFN-γ responses, adjusting for age, graft function and mycophenolate mofetil use. In conclusion, both antibody and T cell responses predict the risk of BTI in kidney transplant recipients who received three doses of SARS-CoV-2 mRNA vaccine. T cell responses may help compensate for the suboptimal antibody response to vaccination in this vulnerable population.

**One Sentence Summary:** Antibody and T cell responses to booster SARS-CoV-2 vaccination predict the risk of symptomatic breakthrough infection in kidney transplant recipients

## INTRODUCTION

Solid organ transplant (SOT) recipients are at high risk of severe COVID-19 and death *(1–4)*. This high risk has been attributed to immunosuppressive therapies and to common co-morbidities, including cardiovascular disease, diabetes, and obesity, that have also been associated with severe COVID-19 and death in the general population *(5, 6)*. While they had not been included in COVID-19 vaccine trials, SOT recipients were offered COVID-19 vaccination early in vaccination campaigns. Unfortunately, their responses to COVID-19 vaccines were weaker than those of healthy adults: neutralizing antibodies were not detectable in most patients, even after two doses of mRNA vaccine *(7)*. These low vaccine responses were associated with a high incidence of breakthrough infections (BTI), including severe cases *(8–12)*. SOT recipients were therefore prioritized for a booster with a third dose of COVID-19 vaccine. Higher levels of SARS-CoV-2 neutralizing antibodies were induced by booster as compared to primary vaccination in these patients, in line with what is observed in the general population *(13–15)*. Yet, the level of immunity against BTI achieved by booster vaccination of SOT recipients, especially in the context of emerging SARS-CoV-2 variants, remains poorly defined *(16)*.

Serum levels of SARS-CoV-2 neutralizing antibodies are correlated with protection against COVID-19 in the general population *(17–20)*. In addition, booster mRNA vaccination is associated with improved vaccine effectiveness in healthy adults *(21)*. An immune correlate of protection against COVID-19 in SOT patients has not been identified. Despite their lower levels in SOT recipients, neutralizing antibodies are likely to play a key role given their importance in other populations. On the other hand, other immune effectors, in particular T lymphocytes, could contribute to protection in patients with limited vaccine-induced humoral immunity. Notably, no clinical study has yet identified a correlation between T cell responses to vaccination and protection against COVID-19 in the general population, possibly because of a dominant role for neutralizing antibodies when induced at high levels *(22)*.

Sociodemographic and clinical factors could also contribute to the risk of BTI in SOT patients. Previous studies identified age, female gender and co-morbidities, such as metabolic syndrome and kidney disease, as risk factors for SARS-CoV-2 BTI *(23, 24)*. It is unclear whether these factors increase the risk of BTI directly, or by reducing the immune response to COVID-19 vaccination. Identifying risk factors for BTI in SOT recipients and their potential impact on COVID-19 vaccine immunogenicity is required to identify high risk patients and to guide COVID-19 booster immunization strategies.

This prospective cohort study was undertaken to identify risk factors of BTI in kidney transplant recipients who had received three doses of BNT162b2 COVID-19 mRNA vaccine. Associations with vaccine-induced immune responses, sociodemographic and clinical factors were explored.

## RESULTS

### Study population

Sixty-six kidney transplant recipients were screened for the study. One patient was excluded because of a positive PCR test on the day of the first COVID-19 vaccination. Characteristics of the 65 patients included in the study are shown in table S1. Mean age was 58.2 years. Mean duration of transplantation was 7.4 years. Immunosuppressive treatment included steroids (78%), mycophenolate mofetil (52%), azathioprine (25%), tacrolimus (68%), cyclosporine (14%) and everolimus (23%). During follow-up, one patient required chronic dialysis, one patient withdrew consent, one patient was lost to follow-up between the second and the third vaccination, and two patients declined the third dose of vaccine (Fig. 1). No patients died during the study period.

**Fig. 1.**
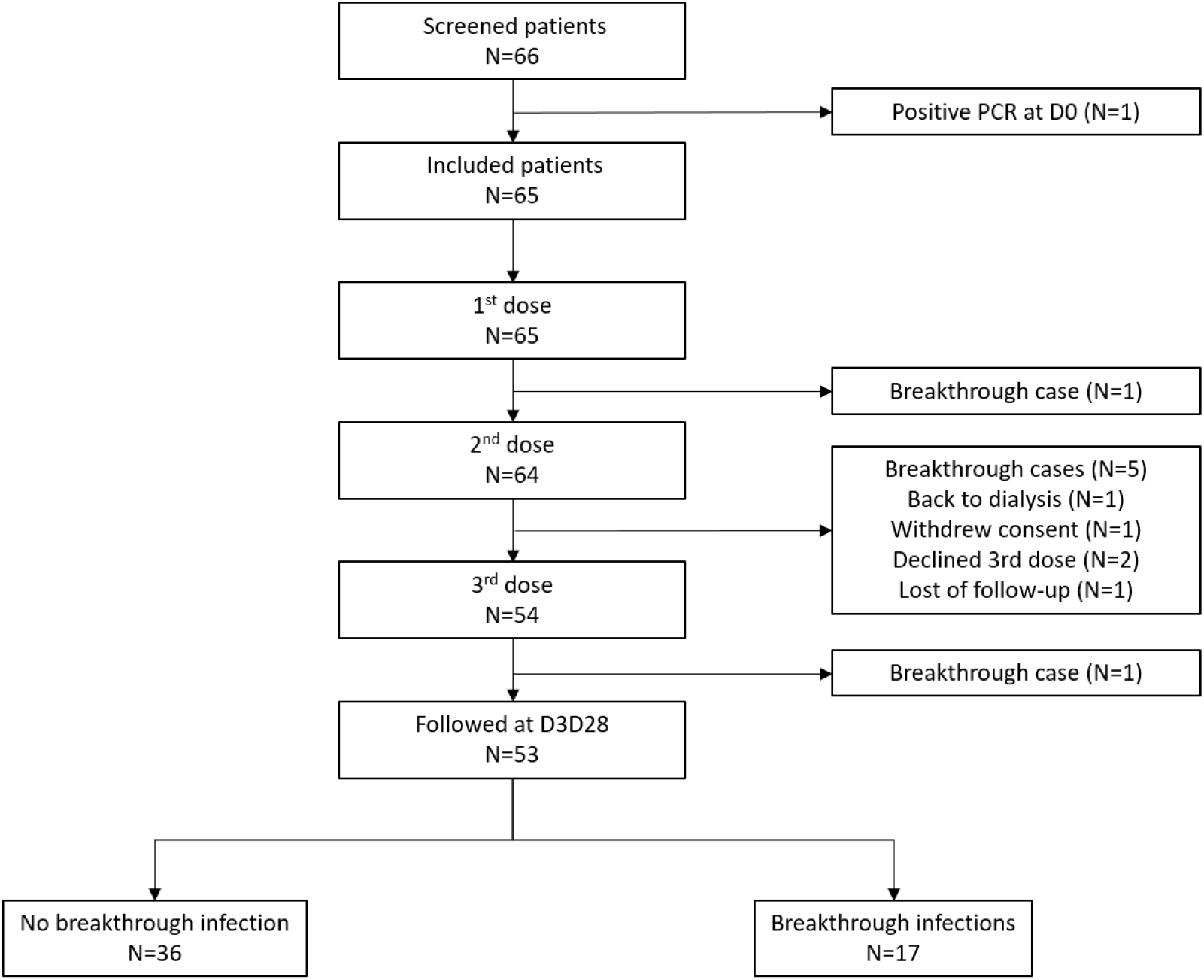
Study flow chart.

### Incidence of SARS-CoV-2 BTI

Six cases of symptomatic SARS-CoV-2 BTI were diagnosed between the second and the third COVID-19 vaccination. Thus, 54 patients were followed-up and received the third dose of vaccine, on average 161 days after having received the first vaccine dose (Fig. 1). One case of symptomatic BTI was diagnosed within 28 days of the third dose of vaccine. At D3D28, 53 patients remained free of symptomatic BTI. Seventeen cases were diagnosed between 28 days after the third vaccination and the fourth vaccination, when the study follow-up ended (Fig. 1). Symptoms related to BTI are shown in table S2. One patient experienced moderate COVID-19 requiring oxygen supply, and the other 16 patients had mild COVID-19. Curative monoclonal antibodies were administered to 14 patients (83%). At the end of the study period, 36 patients remained free of symptomatic BTI.

Figure 2 depicts the cumulative number of symptomatic BTI in the study population over time and in relation to the incidence of SARS-CoV-2 infection with individual variants of concern in the Belgian population. During the first period (before D3D28), the incidence rate of SARS-CoV-2 BTI in the study population was 0.63 (0.25; 1.30) cases per 1000 person-days. During this period, the daily incidence of COVID-19 was estimated at less than 1 case per 1000 inhabitants in Belgium, and the alpha and delta VOC were dominant. During the second period (after D3D28), the incidence rate of BTI in the study population was about 4-fold higher: 2.74 (1.60; 4.38) cases per 1000 persons-days (comparison of incidence rates between the two periods: p<0.001). During this second period, the estimated incidence of COVID-19 also increased in the Belgian population and the delta and BA.1 omicron VOC were dominant (Fig. 2).

**Fig. 2.**
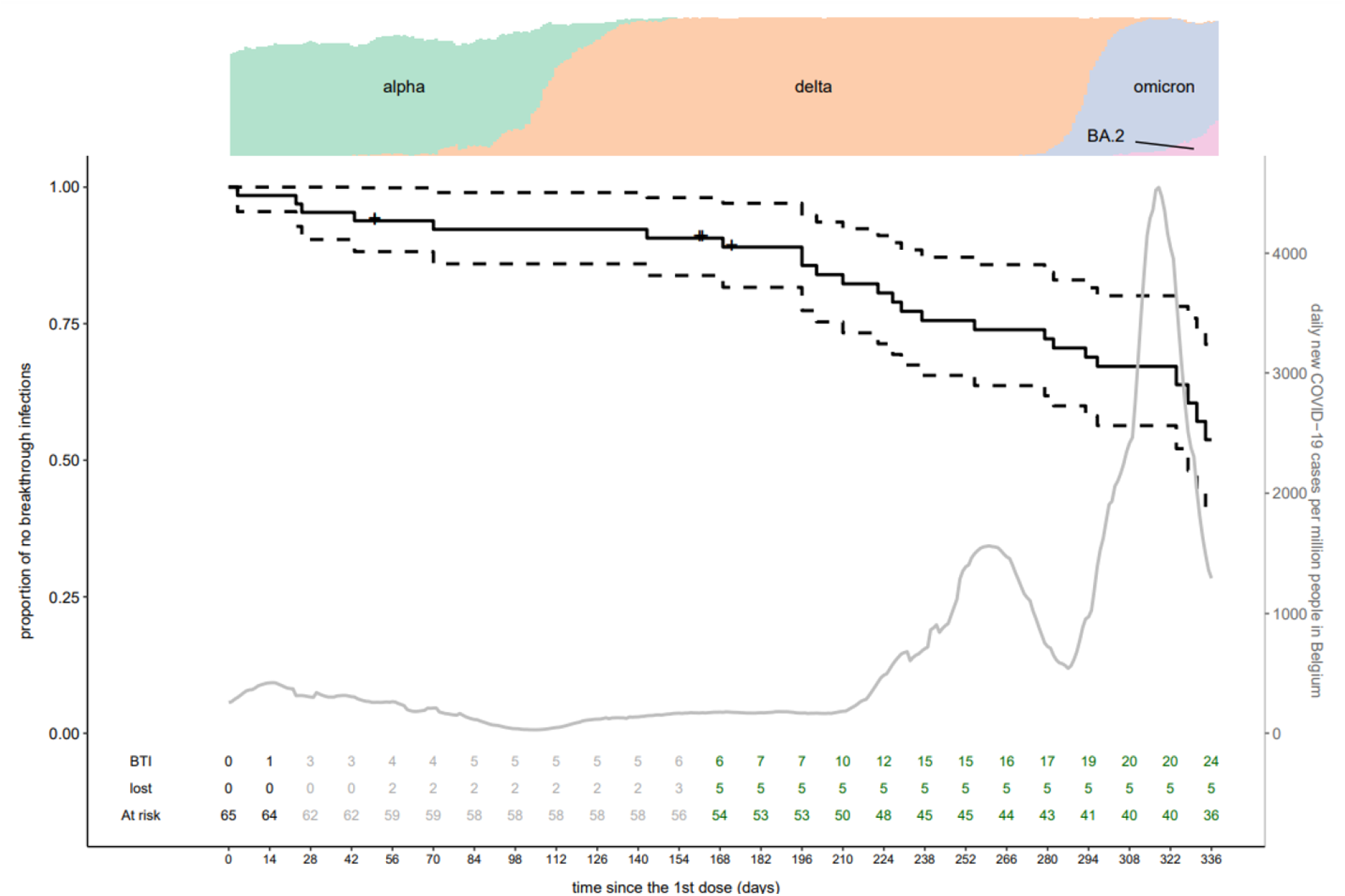
Occurrence of symptomatic breakthrough infection in relation to incidence of COVID-19 in the Belgian population. The black line represents the proportion of kidney transplant recipients who remained free of SARS-CoV-2 BTI during the study period. Dotted lines are 95% confidence intervals. Day 0 is the date of the first vaccine dose for each patient. Patients were followed-up until the first day of administration of the 4^th^ vaccine dose (February 1^st^, 2022). Each cross represents a patient lost to follow-up. Numbers of patients with BTI, lost to follow-up and at risk of BTI are indicated in the lower part of the graph (black numbers: before the second vaccine dose; grey: between the second and third dose of vaccine; green: after the third vaccine dose). The grey line represents daily confirmed COVID-19 cases per million people 7 days rolling average in Belgium. Day 0 for the grey line is the median date of first vaccination of the study population (March 10^th^, 2021). Proportions of infections with VOC are presented in the upper part of the figure (alpha in green, delta in orange, omicron BA.1 in grey and omicron BA.2 in pink, and others in white).

### Immune response to COVID-19 vaccination

Administration of a third dose of COVID-19 vaccine significantly increased the levels and quality of SARS-CoV-2 spike-specific antibodies (Fig. 3 and table S3). Geometric mean serum concentration of RBD binding IgG increased from 8 IU/mL after two doses of vaccine to 35 IU/mL after three vaccine doses. The proportion of patients with detectable RBD binding IgG was 51% after three vaccinations (27/53). Avidity of RBD binding IgG also increased in patients with detectable RBD IgG with a five-fold slower dissociation rate between the second and third dose of vaccine (from geometric mean of k_off_ : 0.0016 s^-1^ after the second dose to geometric mean of k_off_ : 0.0003 s^-1^ after the third dose of vaccine). Neutralizing antibody levels increased similarly, with only 2% of patients having detectable SARS-CoV-2 neutralizing antibodies after two doses and 45% of patients after three vaccine doses. In contrast, cellular immune responses to SARS-CoV-2 were similar after the second and third doses of vaccine (Fig. 3 and table S3).

**Fig. 3.**
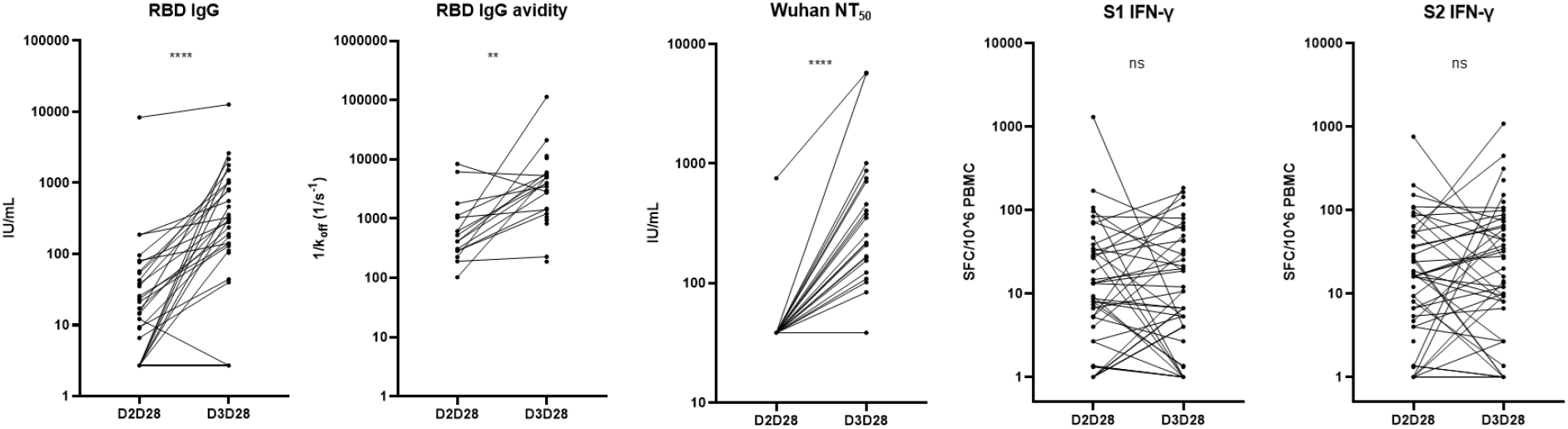
Immune response to second and third COVID-19 mRNA vaccinations in kidney transplant recipients. Immune responses, including SARS-CoV-2 receptor binding domain (RBD)-specific binding IgG titer (RBD IgG) and avidity (RBD IgG avidity), SARS-CoV-2 Wuhan neutralizing antibody titer (Wuhan NT_50_) and S1 (S1 IFN-γ) and S2 (S2 IFN-γ) domain-specific IFN-γ-producing cells, were measured one month after the 2^nd^ dose (D2D28) and one month after the 3^rd^ dose (D3D28) of vaccine among kidney transplant recipients naive of symptomatic BTI at D3D28 (n=53). Each data point represents a sample. Samples were available for 51 patients at D2D28 and 53 patients at D3D28. The lower limit of detection (LLOD) is 5.4 IU/mL for RBD binding IgG and 77 IU/mL for Wuhan NT_50_. Twenty-one patients had detectable RBD binding IgG at D2D28 and 27 at D3D28. One patient had detectable neutralizing antibodies at D2D28 and 24 at D3D28. RBD IgG avidity was only measured for samples with RBD binding IgG levels > 5.4 IU/mL. Statistical comparisons were made using Wilcoxon signed rank test. RBD: receptor binding domain. Wuhan NT_50_: 50% neutralizing antibody titer against Wuhan strain. S1 or S2 IFN-γ: S1 or S2 specific PMBC responses. ** p<0.01; and **** p<0.0001.

### Risk factors of SARS-CoV-2 BTI following booster COVID-19 vaccination

Potential risk factors for SARS-CoV-2 BTI after the third dose of COVID-19 vaccine were first explored by univariate analysis using logistic regression (Fig. 4 and table S4). An increased risk of BTI in kidney transplant recipients was associated with low RBD IgG avidity (odds ratio (95% CI): 0.07 (0.003; 0.61), p=0.04), low titer of neutralizing antibodies (0.15 (0.02; 0.64), p=0.027), and low frequency of S2-specific, and not S1-specific, IFNγ-producing cells (0.30 (0.11; 0.69), p=0.009). No significant association was detected between the risk of BTI following booster COVID-19 vaccination and the demographic or clinical variables that were analyzed (Fig. 4 and table S4).

**Fig. 4.**
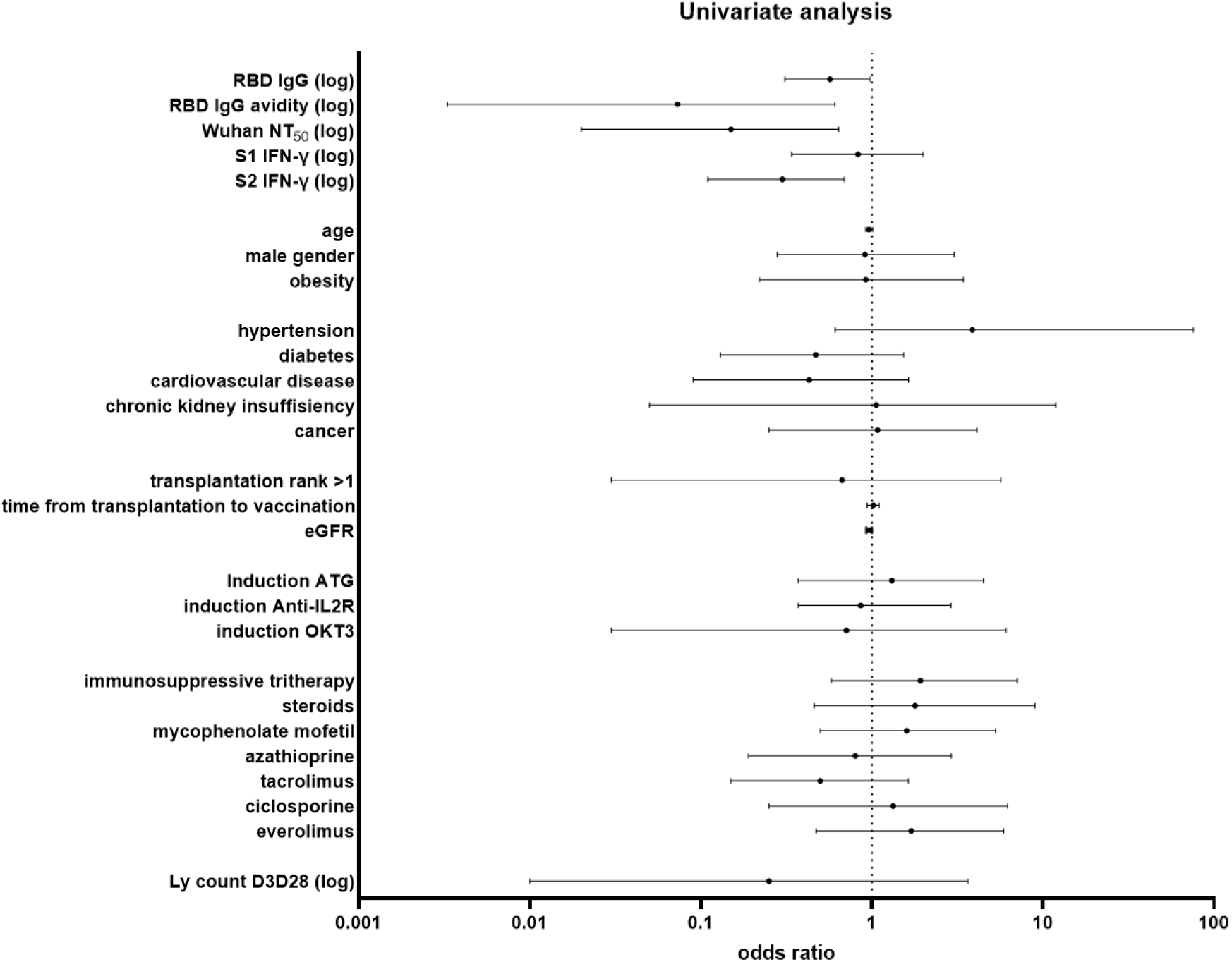
Univariate analysis of risk factors of symptomatic breakthrough infection in kidney transplant recipients. Univariate logistic regression was used to assess the relationship between immunological, demographic and clinical parameters and the occurrence of symptomatic BTI. Obesity was defined as BMI > 30 kg/m^2^. Transplantation rank was divided into first transplantation and second or more transplantation. Immunological parameters and absolute lymphocyte counts were log10 transformed before analysis. RBD: receptor binding domain. Wuhan NT_50_: 50% neutralizing antibody titer against Wuhan strain. S1 or S2 IFN-γ: S1 or S2-specific PMBC responses. eGFR: estimated glomerular function rate. OKT3: muromumab; ATG: anti-thymoglobulin. Ly count: absolute lymphocyte count.

The absence of significant association between demographic and clinical variables and the risk of BTI led us to explore their potential association with the immune response to COVID-19 vaccination. As shown in table S5, older age was associated with lower humoral immune responses to booster vaccination. Lower kidney graft function and mycophenolate mofetil use were associated with lower humoral and cellular immune responses to booster vaccination, whereas the use of azathioprine was associated with higher vaccine responses. Together, these results indicate that demographic and clinical variables are associated with differences in immune responses to booster COVID-19 vaccination in kidney transplant recipients even if they are not significantly associated with the risk of BTI in this population.

### Multi-parametric analysis of immune response to booster COVID vaccination and the risk of BTI

Multi-parametric serology analyzes were performed to further explore the association between immune response to booster vaccination and the risk of BTI in kidney transplant recipients. In multi-parametric analysis, patients who developed SARS-CoV-2 BTI after booster vaccination had significantly lower RBD binding IgG titer and lower levels of neutralizing antibodies as compared to patients who remained free of symptomatic BTI (Fig. 5A). A similar same trend was observed with RBD IgG avidity. Patients with BTI also had lower levels of S1-specific, but not S2-specific, binding IgG (Fig. 5B). In parallel, patients with BTI had lower levels of S1-specific, but not S2-specific, IgG activating cellular phagocytosis (antibody-dependent cellular phagocytosis, ADCP). After booster vaccination, only few patients had detectable neutralizing antibodies against delta (n=4), omicron BA.1 (n=1) and omicron BA.2 (n=4) (table S6). None of them developed symptomatic BTI. Analysis of cellular immune responses confirmed the results of the univariate analysis (Fig. 4): patients with BTI had significantly lower frequencies of S2-specific, and not S1-specific, IFN-γ producing cells, as compared to patients with no BTI (Fig. 5C). Humoral immune response parameters (RBD IgG, RBD IgG avidity, Wuhan NT_50_, S1 IgG, S2 IgG, S1 IgG ADCP and S2 IgG ADCP) were highly correlated (Fig. 5D). In contrast, significant but lower correlations were observed between humoral and cellular immune response (S1 IFN-γ and S2 IFN-γ) parameters.

**Fig. 5.**
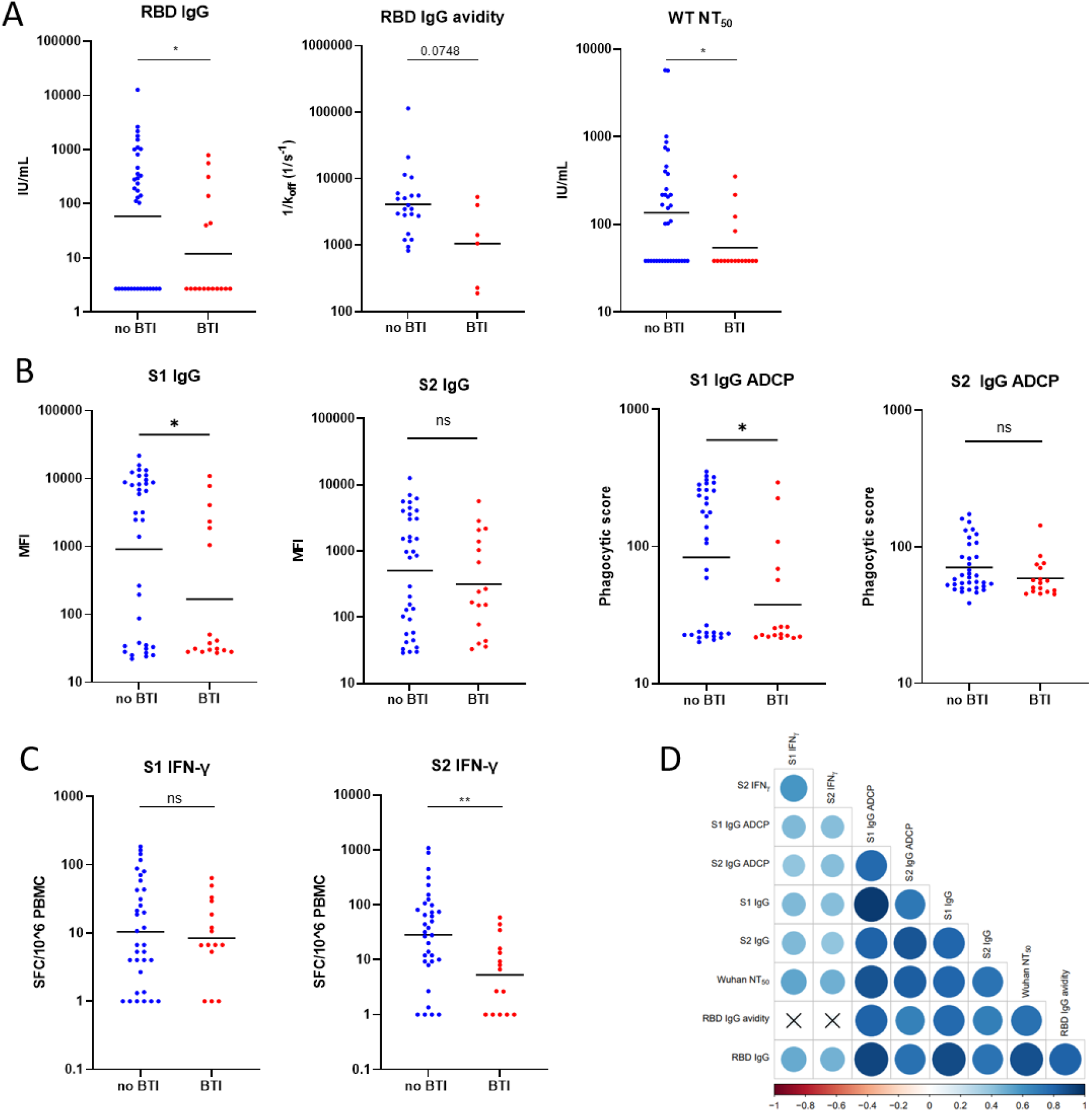
Immune response to booster COVID-19 vaccination in kidney transplant recipients with and without symptomatic BTI. Humoral and cellular immune responses were measured 28 days after a third dose of BNT162b2 COVID-19 vaccine in 53 kidney transplant recipients who developed (red symbols) or did not develop (blue symbols) symptomatic SARS-CoV-2 BTI following vaccination. Black bars indicate geometric means. Variables were compared with Wilcoxon-Mann-Whitney test. * p<0.05; ** p<0.01. **(A)** SARS-CoV-2 receptor binding domain (RBD)-specific binding IgG titer (RBD IgG) and avidity (RBD IgG avidity), SARS-CoV-2 Wuhan neutralizing antibody titer (Wuhan NT_50_). **(B)** S1-specific binding IgG (S1 IgG), S2-specific binding IgG (S2 IgG), S1-specific binding IgG antibody –dependant cellular phagocytosis (S1 IgG ADCP) and S2-specific binding IgG antibody –dependant cellular phagocytosis (S1 IgG ADCP). **(C)** S1-specific cells producing IFN-γ (S1 IFN-γ) and S2-specific cells producing IFN-γ (S2 IFN-γ). **(D)** Pearson correlation matrix of immune response parameters. Correlation coefficients are color coded. Circle size indicates the absolute value of corresponding correlation coefficients. All correlations were statistically significant, except those marked by a cross instead of a circle.

### Multivariate analysis of risk factors of BTI

Multivariate analysis was performed to identify the immunological factors that best predicted the risk of BTI following booster vaccination. Based on the associations observed between clinical variables and COVID-19 vaccine responses (table S5), this analysis was adjusted for age, kidney graft function (eGFR) and mycophenolate use. Only variables with a variance inflation factor (VIF) <10 (RBD binding IgG, Wuhan NT_50_, S1 IgG, S2 IFN-γ) were included in the analysis. Wuhan neutralizing antibody titer and S2 IFN-γ responses were the best predictors of BTI following booster vaccination, with odds ratios of 0.036 ((<0.001; 0.501), p=0.028) and 0.145 ((0.016; 0.671), p=0.037), respectively. Based on this analysis, we estimated that patients with a 10-fold increase in Wuhan NT50 had on average a 28-fold lower risk of BTI and patients with a 10-fold increase in S2 IFN-γ responses had on average a 7-fold lower risk of BTI.

## DISCUSSION

In this prospective cohort study, we observed that both humoral and cellular immune responses to booster COVID-19 vaccination predict the risk of symptomatic BTI in kidney transplant recipients. Although age, kidney graft function and the use of mycophenolate were associated with altered vaccine responses, no significant association was observed between these parameters, or other demographic and clinical parameters, and the occurrence of BTI. These data emphasize the importance of inducing adequate immunity through vaccination to protect this vulnerable population from COVID-19.

Our analysis was focused on kidney transplant recipients who had not been diagnosed with COVID-19 either before vaccination or before the response to booster vaccination was measured. Patients had low humoral immune responses after two doses of mRNA vaccine, confirming previous studies in SOT recipients *(13–15)*. Humoral immunity was significantly enhanced by booster vaccination, as observed in the general population *(21)*. In contrast, cellular immune responses were similar following two doses of vaccination and after booster vaccination. Enhanced cellular immune responses following booster vaccination was observed in the general population, suggesting that boosting of cellular immunity may be more difficult to achieve in SOT recipients as compared to healthy adults *(25)*. Despite enhanced immunity following booster vaccination, patients remained at high risk of SARS-CoV-2 BTI. Thirty two percent of our study population developed symptomatic BTI in the first 6 months following booster vaccination. This incidence was significantly higher than the one observed before booster vaccination, most likely because of the higher virus circulation of delta and omicron VOC during this period. This observation further emphasizes the importance of booster vaccination to prevent COVID-19 in SOT recipients *(8, 9)*. Most patients received therapeutic monoclonal antibodies to prevent severe COVID-19 and only one patient required oxygen supplementation.

To our knowledge, this is the first report on correlate of protection against COVID-19 in SOT recipients. As observed in the general population, the antibody response to COVID-19 vaccination was associated with protection of kidney transplant recipients against BTI with VOC *(20)*. As already published in the general population, the strongest association with protection against BTI in SOT was observed with Wuhan neutralizing antibody titer *(18, 20)*. In addition to SARS-CoV-2 neutralization, antibodies mediate viral control via Fc-dependent effector functions, including ADCP. We observed high correlations between the titers of binding, neutralizing and ADCP-promoting antibodies against SARS-CoV-2 RBD and S1 subunit, indicating a strong coordination between the diverse effector components of the antibody response. S1-specific ADCP-promoting IgG were higher in patients who remained free of BTI following booster vaccination, supporting the notion that they may also contribute to vaccine-induced protection. In contrast, S2-specific binding and ADCP-promoting IgG were similar in patients who developed or did not develop BTI. This observation suggests a dominant role of S1-specific antibodies in mediating protection against BTI in this population.

The S2-specific IFN-γ response to booster vaccination was the second most important predictor of BTI. As no significant association was observed with S1-specific IFN-γ responses, our data suggest a differential role of SARS-CoV-2 subunits in mediating protection following mRNA vaccination. As the S2 subunit is more conserved than the S1 subunit across SARS-CoV-2 VOC, it may be an important target for cell-mediated immunity against the delta and omicron variants to which our study population was exposed. To our knowledge, this is the first study indicating a role for cellular immune response to vaccination in protection against COVID-19. As suggested by Kent et al, a role for T cell immunity may be difficult to reveal in healthy adults who have optimal antibody responses to vaccination that may dominate protective immunity against COVID-19 *(22)*. The suboptimal humoral immune response to vaccination observed in SOT recipients may increase the importance of cellular immune responses in this population. Supporting this notion, a role for vaccine-induced T cell immunity in the context of low humoral immunity was shown in non-human primate studies of COVID-19 *(26)*.

This study has several limitations. First, its sample size is relatively low because it was a single centre study. This limitation was decreased by the high incidence of symptomatic BTI during a period when delta and omicron VOC were dominant, allowing us to detect statistically significant associations with immunological parameters. A larger sample size may have allowed us to detect significant associations with demographic and clinical parameters. Further studies of correlates of protection in SOT recipients are needed to validate our observations. Second, the study did not include systematic screening of SARS-CoV-2 by PCR and therefore did not allow us to diagnose asymptomatic SARS-CoV-2 BTI. Such systematic screening was considered too challenging by the patients and by the study team. Instead, priority was placed on diagnosing symptomatic BTI, which we consider to be diagnosed with high sensitivity. Third, the study does not provide information regarding immunity against severe COVID-19, as most patients experiencing a BTI received therapeutic monoclonal antibodies and consequently presented mostly mild symptoms. Finally, while the very strong dominance of delta and omicron VOC in the Belgian population during the study period indicates that these were likely the cause of BTI in our study population, sequencing of SARS-CoV-2 was not performed systematically following diagnosis of BTI. Differences in correlates of protection against delta and omicron variants have been suggested and may have influenced our results *(27)*.

In conclusion, this study shows that humoral and cellular immune responses induced by booster vaccination correlate with protection against SARS-CoV-2 BTI in kidney transplant recipients. These data emphasize the importance of reaching and maintaining a high level of immunity through vaccination in this vulnerable population and suggest that vaccines inducing potent cellular immune responses, such as mRNA vaccines, may be particularly useful in populations with suboptimal humoral immune responses to vaccination.

## MATERIALS AND METHODS

### Study design and population

This single centre prospective phase IV investigator-initiated study of the immunogenicity of the BNT162b2 vaccine (Pfizer-BioNTech) in kidney transplant recipients was approved by the ethics committee of the Erasme Hospital (P2020/284 and A2021/131) and by the Belgian Federal Agency for Medicines and Health Products (FAMHP, EudraCT 2021-000-412-28). Kidney transplant recipients aged at least 18 years were recruited in the Department of Nephrology, Dialysis and Transplantation of the Erasme Hospital, Belgium. They were informed and gave informed consent before administration of the first dose of BNT162b2 vaccine. Patients transplanted with multiple organs were excluded from the study. Patients infected with SARS-CoV-2 before BNT162b2 vaccination were not included in this report. Previous SARS-CoV-2 infection was defined as a history of a positive PCR on nasopharyngeal swab before study recruitment and/or a positive serology (Wantai SARS-CoV-2 IgG Elisa, see Supplementary methods) at recruitment, before administration of the first dose of BNT162b2 vaccine. Patients were seen by a study clinician at enrolment before the first dose, one month after the second dose and the day of the third dose of vaccine. Demographic and clinical data were recorded, including age, gender, body mass index, comorbidities (hypertension, diabetes, cardiovascular disease, chronic kidney insufficiency defined with eGFR < 30mL/min in CKD-EPi and evolutive or treated cancer), characteristics of transplantation history (duration, rank, graft function, induction treatment and immunosuppressive drugs), and absolute lymphocyte count.

All patients received the BNT162b2 vaccine at day 0 and day 21, according to the Belgian national vaccination program. The first vaccine dose was administered between March 2^nd^ and March 18^th^, 2021, and the second dose between March 23^rd^ and April 8^th^, 2021. The third dose of BNT162b2 vaccine was administered between August 17^th^ and 20^th^, 2021, following an amendment of the study protocol approved by the Belgian FAMHP. BNT162b2 is a lipid nanoparticle–formulated, nucleoside-modified mRNA vaccine encoding a prefusion stabilized, membrane-anchored SARS-CoV-2 full-length spike protein *(28)*. The 30μg/0.3 mL dose was administered by intra-muscular injection into the upper arm. Vaccines were administered by a trained study nurse at Erasme Hospital. Participants were observed for a minimum of 15 minutes after vaccination. No serious adverse events were observed.

### Breakthrough infection

Following recruitment, symptomatic SARS-CoV-2 breakthrough infections (BTI), defined as the occurrence of fever, chills, cough, dyspnoea, rhinitis, anosmia, dysgeusia, abdominal pain or diarrhoea, associated with a positive SARS-CoV-2 PCR on nasopharyngeal swab, were recorded. Patients were informed of symptoms compatible with COVID-19 and were encouraged to be tested by PCR of a nasopharyngeal swab in case of such symptoms. Patients were regularly contacted by phone and/or by email to limit unreported BTI. The following data were recorded when a BTI was diagnosed: symptoms, date of symptom onset, oxygen requirement, organ failure and treatment, including curative monoclonal antibodies. The date of BTI was defined as the date of symptom onset. Only the first symptomatic BTI was included in the data analysis.

### Immune response to BNT162b2 vaccination

Humoral and cellular immune responses to BNT162b2 vaccination were measured one month after the second vaccine dose (D2D28) and one month after the third vaccine dose (D3D28). Pre-vaccination immune responses to SARS-CoV-2 were analyzed as baseline. Humoral immune responses measured included SARS-CoV-2 receptor binding domain (RBD)-specific binding IgG and avidity, SARS-CoV-2 Wuhan, B.1.617.2 Delta variant (83DJ-1), BA.1 Omicron variant and BA.2 Omicron variant neutralizing antibodies, and SARS-CoV-2 S1 and S2 domain binding and phagocytosis inducing IgG. Cellular immune responses measured included S1 and S2 domain-specific IFN-γ-producing cells. For detailed laboratory methods, see Supplementary Material.

### Statistical analyzes

Occurrence of SARS-CoV-2 BTI during the study period (from March 2^nd^-18^th^, 2021, to February 1^st^, 2022) was plotted as a Kaplan-Meier survival curve. The incidence of SARS-CoV-2 infection diagnosed in Belgium during the study period and the proportion of infections with variants of concern (VOC) circulating in Belgium were included in the graph to provide the epidemiological context of the study *(29)*. Incidence rate (person-day rate) of BTI was defined as the ratio of breakthrough infections (BTI) divided by the time each patient was observed, totalled for all the patients. It was calculated for two different periods: (1) from the first day of vaccination to D3D28, and (2) from D3D28 to February 1^st^, 2022, when the fourth dose of COVID-19 vaccine was administered. Descriptive data were evaluated with geometric means (95% CI) for continuous variables, and n (%, 95% CI) for binary values. Descriptive data were compared by *t*-test or Wilcoxon-Mann-Whitney rank test for parametric and non-parametric values, respectively, according to D’Agostino normality test. A Wilcoxon signed rank test was used to test paired sampled, when comparing two time points. The proportion of participants with antibody levels higher than the lower limit of detection (LLOD) at D2D28 or D3D28 were calculated for SARS-CoV-2 RBD binding IgG (LLOD: 5.4 IU/mL according to WHO standard) and neutralizing antibodies (LLOD: 77 IU/mL according to WHO standard). For RBD binding IgG and neutralizing antibodies, antibody concentrations below the limit of detection were assigned a value equal to half of the LLOD before transformation. RBD IgG avidity was measured for samples with detectable binding RBD IgG only. For IFN-γ ELISpot, data equal to 0 or less than 0 after subtraction of background were assigned a value of 1 before transformation. Values higher than 300 spots forming cells/250,000 PBMC were censured at 1.200/ 10^6^ PBMC. The proportion of participants with cellular responses higher than an arbitrary cut-off of 50 SFC/10^6 PBMC in IFN-γ ELISpot were calculated at D2D28 or D3D28. Association between immune variables and demographic and clinical variables were explored by linear regressions. Univariate logistic regression was used to explore the factors associated with the occurrence of BTI for the period between D3D28 and February 1^st^, 2022. Variables with a p-value < 0.1 in univariate analyzes were included in a variance inflation factor (VIF) analysis. To avoid multi-collinearity, only variables with VIF<10 were included in multivariate analysis. A multivariate logistic regression model was built to analyze risk factors of BTI, adjusting for age, kidney graft function and mycophenolate mofetil use, considered as important variables influencing immune responses to SARS-CoV-2 vaccination, based on our linear regression data and on published data *(30, 31)*. Missing data were excluded from the final analysis. The final model was selected based on backward selection of variables. All statistical analyzes were done using R version 4.2.0 *(32)*.

## Data Availability

All data produced in the present study are available upon reasonable request to the authors

## Supplementary Materials

### Supplementary illustrations

table S1. Demographic and clinical characteristics of naive kidney recipients at baseline

table S2. Characteristics of symptomatic SARS-CoV-2 BTI

table S3. Humoral and cellular immune responses at D2D28 and D3D28.

table S4. Univariate analysis assessing the relation between breakthrough infections and clinical or immunological variables

table S5. Linear regression assessing the association between immune parameters and demographical or clinical variables.

table S6. Binding and neutralizing antibody titers by variant of concern after three vaccine doses (D3D28).

## General

We thank Sorya Fagnoul and Emilie Navaux for the help in the recruitment and follow-up of patients. Maria Spanu and Anne Gillardin for the sampling and the follow-up of patients. Inès Vu Duc, Florian Ballieu, Vianney Nana Tchouta, Marie-Annick Ottou Eyenga, Sara Cuman, Robin Van Naemen and Caroline Rodeghiero for technical assistance. We thank the Robotein^®^ platform of the BE Instruct-ERIC Centre for providing access to the Octet HTX and the Microlab STAR liquid handling workstation. Finally, we thank all patients for their generous participation in the study.

## Funding

This study was co-funded by the Belgian Federal Government through Sciensano and by the Fonds de la Recherche Scientifique, F.R.S.-FNRS, Belgium.

D.K. received a PhD studentship from the F.R.S-FNRS, Belgium.

N.G. received a PhD studentship from the Fonds Erasme, Belgium.

A.M. is Research Director of the F.R.S.-FNRS, Belgium.

## Author Contributions

DK, NG, ALM and AM conceptualized the study. DK and PP wrote the clinical study protocol. DK obtained permission from the ethics committee and AFMPS. MEG, ALM and AM secured the funding of the study. DK, NG and AL conducted the clinical study. ALM overviewed the clinical study. DK, NG, VO, DG, MV, AW, JM and LH conducted the laboratory analyzes. AM, PP, ID, MEA, AMat and KKA overviewed the laboratory analyzes. DK, NG, SD, DG and LH analyzed the data. DK, NG, SD, MEA and AM interpreted the data. DK and AM drafted the manuscript. All co-authors reviewed and approved the manuscript.

## Data and materials availability

The datasets used and analyzed during the current study are available from the corresponding author on reasonable request, following materials transfer agreements.

