## Supplementary Materials for "Humoral and cellular immune correlates of protection against COVID-19 in kidney transplant recipients"

### Supplementary methods

#### Clinical sample processing and storage

Serum was stored at -80°C before testing. PBMC (peripheral blood mononuclear cells) were isolated from heparinized whole blood by density gradient centrifugation following the recommended protocol (Leucosep tubes, Greiner). Whole blood was diluted with an equal amount of medium. Upper plasma layer was discarded without disturbing the interface. PBMC layer was then retained and washed three times with medium. Cell pellet was then diluted with FBS – 10% DMSO. PBMC were frozen and stored in liquid nitrogen. The day of the T-cell response analysis, PBMC had a one-hour rest before counting, after thawing.

#### Enzyme-linked immunosorbent assay

Serum levels of RBD binding IgG were measured using an enzyme-linked immunosorbent assay (ELISA) (Receptor Binding Domain, Wuhan strain) (Wantai SARS-CoV-2 IgG ELISA (Quantitative); CE-marked; WS-1396; Beijing Wantai Biological Pharmacy Enterprise Co., Ltd, China). Diluted serum samples (1/10, 1/100, 1/400, 1/1600 and 1/6400) were tested with an internal standard, calibrated against NIBSC 20/136 (First WHO International Standard Anti-SARSCoV-2 Immunoglobulin), and an external positive control sample included on each plate. Diluted samples were incubated (37°C, 30 min.) with pre-coated micro wells and washed five times. Next, plates were incubated (37°C, 30 min) with horseradish peroxidase (HRP)-conjugated anti-human IgG antibodies and washed five times before adding a TMB and urea peroxide solution for 15 min (37°C, dark). After incubation, a stop solution (0.5 M H2SO4) was added, and optical density (OD) was measured at 450 nm using a microplate reader. Net OD values were converted to arbitrary IgG units per ml by interpolation from a point-by-point plot fitted with the standard concentrations and net OD values (correlation coefficient $R^2 \geq 0.9801$), using GraphPad Prism version 9.0.0 for Windows (GraphPad Software, San Diego, California USA) and exported to Microsoft Excel. Antibody measurements were adjusted for sample dilution, converted to international units per ml (IU/ml), and reported as such. Lower limit of detection (LLOD) was 5.4 IU/ml, according to WHO standard. Clinical performance characteristics of the assay, evaluated in 69 PCR-confirmed COVID-19 patients (comprising mild and severe clinical outcomes, ≥ 15 days post onset of symptoms) and 167 pre-pandemic sera, resulted in a specificity of 100% (95% CI 97.75-100) at a sensitivity of 100% (95% CI 94.73-100) for a cut-off of 5.4 IU/ml. Data lower than the limit of detection were attributed the value 2.7 IU/mL before transformation.

#### SARS-CoV-2 Neutralizing Antibodies

Serial dilutions of heat-inactivated serum (1/50-1/25600 in EMEM supplemented with 2mM L-glutamine, 100U/ml - 100μg/ml of Penicillin-Streptomycin and 2% foetal bovine serum) were incubated during 1h (37°C, 7% CO2) with 3xTCID100 of a wild type Wuhan strain (2019-nCoV-Italy-INMI1, reference 008V-03893), the B.1.617.2 Delta variant (83DJ-1) and the BA.1 Omicron variant and BA.2 Omicron variant of SARS-CoV2 in parallel. Sample-virus mixtures and virus/cell controls were added to Vero cells (18.000 cells/well) in a 96-well plate and incubated for five days (37°C, 7% CO2). The cytopathic effect caused by viral growth was scored microscopically. For quantification of neutralizing antibodies serum samples were tested with an internal standard, calibrated against NIBSC 21/234 (WHO International Standard Anti-SARS-CoV-2 Immunoglobulin). The Reed-Muench method was used to calculate the neutralizing antibody titer that reduced the number of infected wells by 50% ($NT_{50}$), which values were used as a proxy for the neutralizing antibody concentration in the sample. Data lower than the limit of detection of 77 IU/mL for $NT_{50}$ were attributed the value of 38.5 IU/mL before transformation, according to WHO standard.

#### RBD IgG Avidity

Avidity was assessed by using Biolayer Interferometry (BLI). BLI measurements were performed with the Fortebio HTX Octet instrument and Fortebio AR2G biosensors. Data analyses were performed using ForteBio Data Analysis 12.0 software. Kinetic assays were performed at 25-30˚C at a sample plate agitation speed of 1000 rpm. The use of AR2G biosensors involves several steps before kinetic measurements. First, a sensor activation step is performed by immersing the biosensors in a solution containing 20 mM 1-ethyl-3-[3-dimethylaminopropyl] carbodiimide hydrochloride (EDC) and 10 mM nhydroxysuccinimide (NHS). Then, 0.05 mg/ml of RBD antigen (2019nCoV RBD protein, Sanyou #PNA004) in 10 mM sodium acetate pH 6 was loaded for 600 seconds onto AR2G sensors. After antigen loading, the biosensors were immersed in a solution of 1 M ethanolamine pH 8.5 to inactivate the remaining sites on the biosensors to prevent possible non-specific interactions. The antigen loaded on AR2G sensors were then immersed in a PBS solution (20 mM pH 7.5) to establish a baseline time curve, and then immersed for 600 seconds in wells containing the analytes (polyclonal IgG antibodies purified from serum with the Melon™ gel purification kit (Thermo Scientific™) at different dilutions (diluted in 20 mM PBS pH 7.5) to achieve association. The dissociation step was performed by replating the antibody-bound sensors for 600 seconds in the wells used to collect the baseline time course. Two negative controls were performed: a first one where the ligand is loaded and not the analyte, to ensure the correct loading of the antigen; and a second control without ligand and in the presence of analyte to ensure that there is no non-specific interaction between IgG and AR2G biosensors. Because the concentration of anti-RBD antibody is unknown in a polyclonal antibody solution, samples containing purified IgG solutions were analyzed at three dilutions (3x, 5x, and 8x). Avidity was only determined for samples with RBD IgG titer upper than the limit of detection (5.4 IU/mL). Kinetic parameters were determined by global fitting of the association and dissociation phases of the binding curves following a 1:1 binding model. Avidity was determined using the dissociation rate constant ($k_{off}$) expressed in $s^{-1}$. For statistical analysis and graphical representation, the $k_{off}$ was reversed, indicating that the lower was the $k_{off}$, the higher was the avidity.

#### Enzyme-linked immunosorbent spot Spike-specific cellular responses

Enzyme-linked immunosorbent spot Spike-specific cellular responses were tested after vaccination using the Human IFN-γ ELISpot kit (3420-2H) from Mabtech (Stockholm, Sweden) following the recommended protocol. Pre-coated Plates (Mabtech 3420-4HPW-10) were washed and blocked with 200µl of Roswell Park Memorial Institute (RPMI) containing 10% foetal bovine serum (FBS) for at least two hours. Next, triplicates of 250 000 PBMC were stimulated in the presence or absence of PepMix SARS-CoV-2 spike glycoprotein peptide pools (SUB1-SUB2, JPT, Berlin, Germany) at 1µg/ml and incubated for 18 hours in a 37°C humidified incubator with 5% CO2. After incubation, the plates were washed and incubated with the human biotinylated IFN-γ detection antibody (1µg/ml) for 2 hours, washed and the streptavidin– Horseradish Peroxidase (streptavidin-HRP) diluted at 1/1000 in PBS-0,5% FBS was added for one hour. 3.3', 5.5'-Tetramethylbenzidine substrate was added for eight minutes at room temperature. Wells were then washed with distilled water and air-dried. Cells stimulated with 1µg/ml Staphylococcal enterotoxin (SEB; Sigma-Aldrich) served as positive control. Spots were counted with an ELISpot reader (AID Autoimmun Diagnostika GmbH, Straßberg, Germany), mean values of triplicates were considered for S1 and S2 and expressed per million PBMCs after subtracting the mean of the triplicates of the unstimulated condition. Outliers, defined as a well with a number of spots higher than 3-fold the mean of the two other wells in the same condition, were excluded from the analysis. Data points equal to 0 were attributed the value 1 before transformation. Reponses were expressed as spot forming cells per million of PBMC. Unstimulated samples had spot counts between 0 and 72 SFC/$10^6$ PBMC.

#### Luminex profiling

Detection of SARS-CoV-2–specific IgG antibodies directed against the S1 domain (S1 Sanyoubio, #PNA002) and the S2 domain (S2 SinoBiological, #40590-V08H1) of the wild-type Spike protein of SARS-CoV-2 and the Delta variant of the Receptor-Binding Domain of SARS-CoV-2 (Proteogenix # PX-COV-P061-10) was performed using a 96-well based customized multiplexed Luminex assay, as previously described[21].-Antigens were coupled by covalent NHS-ester linkages via EDC and NHS (Pierce #77149 and #24520, respectively) to fluorescent carboxyl modified microspheres (Luminex). Antigen-coupled microspheres were then washed using magnetic separation and incubated 2 hours at room temperature with plasma samples at appropriate dilution: 1:100, 1:1000 and 1:5000 for IgG titration. Antigen-specific antibodies were detected using 0.65 µg/ml of PE-coupled detection antibody for IgG (Southern Biotech # 2040-09). Antigen-antibody reactions were read with a BioPlex-200 (Bio-Rad) and results were expressed as median fluorescence intensity. Serum from recovered COVID-19 patients were used as positive controls.

#### Phagocytosis Assays

Antibody-dependent cellular phagocytosis (ADCP) was assessed using the human monocyte cell line THP-1 (ATCC #TIB-202). Prior the assay, 0.25 mg/ml of the S1 and S2 of SARS-CoV-2 were biotinylated using EZ-Link NHS-LC- LC-Biotin (Thermo #21343) for 30 min at 37°C (1 mole of antigen for 50 moles of biotin). The biotinylated antigens were then purified using zeba spin desalting columns (Thermo #89883) and then coupled to FluoSpheres NeutrAvidin beads (Thermo #F8776) for 2 h at 37°C. Antigen-conjugated beads were then washed twice with 0.1% BSA in PBS and diluted 100-fold in this buffer. In a 96-well clear microplate (Greiner #650180), 10 ml of antigen-conjugated beads were incubated for 2h at 37°C with 10 ml of diluted purified IgG (1:100). After incubation, wells were washed twice with PBS and centrifugated (2000 g, 10 minutes). 200 ml of 0.5x105 THP-1 cell/ml were added to each well and incubated 16 hours at 37°C, 5% C02. Finally, cells were centrifuged (125 g, 10 min) and 100 ml of the supernatant was substituted by 100 ml of 1X CellFIX (BD #340181). Cells were analyzed using an LSR-Fortessa (BD). Results were expressed as phagocytic score: FITC+frequency X FITC+mean*10^4. Assays performed without IgG and with a pool of IgG obtained from low responders for each antigen were used as negative controls.

#### Supplementary illustrations

| | ALL<br>N=65 |
|---|---|
| **Demographic data** | |
| Age (years) | 58.2 (54.7; 61.9) |
| Male gender (%) | 36 (55) |
| Body mass index (kg/m²) | 25.3 (24.0; 26.6) |
| **Comorbidities** | |
| Hypertension (%) | 55 (85) |
| Diabetes (%) | 24 (37) |
| Cardiovascular disease (%) | 17 (26) |
| Chronic kidney insufficiency (%) | 7 (11) |
| Pulmonary disease (%) | 5 (8) |
| Cancer (%) | 16 (25) |
| **Transplantation data** | |
| Transplantation rank >1 (%) | 7 (11) |
| Time from transplantation to vaccination (years) | 7.4 (5.7; 9.5) |
| eGFR – CKD-EPI (mL/min) | 47.5 (43.2; 52.2) |
| Anti-IL2 receptor (%) | 39 (60) |
| Anti-thymocyte (%) | 19 (29) |
| OKT3 (%) | 5 (8) |
| Unknown induction treatment | 2 (3) |
| Triple immunosuppressive tritherapy (%) | 39 (60) |
| Steroids (%) | 51 (78) |
| Mycophenolate mofetil (%) | 34 (52) |
| Azathioprine (%) | 16 (25) |
| Tacrolimus (%) | 44 (68) |
| Ciclosporine (%) | 9 (14) |
| Everolimus (%) | 15 (23) |
| Absolute lymphocyte count (/µL) | 1220 (1066; 1397) |

##### table S1. Demographic and clinical characteristics of naive kidney recipients at baseline.

Sixty-five patients were included in the prospective cohort study at D0. Data are geometric means (95% CI) for continuous variables and n (%) for binary variables. Hypertension was defined as chronic blood pressure higher than 140/90 or need for chronic treatment. Diabetes was defined as the need for hypoglycemic treatment and a previous Hb1Ac > 6.5%. Cardiovascular disease was defined as a heart insufficiency or ischemic, valvular or rhythmic heart disease or previous ischemic stroke or symptomatic peripheral vascular disease. Chronic kidney insufficiency was defined as an eGFR < 30mL/min. Pulmonary disease was defined as a chronic respiratory insufficiency or the need for chronic treatment. Cancer includes evolutive cancer or cancer in remission for less than five years. eGFR – CKD-EPI: estimated glomerular function rate based on CKD-EPI equation (2021). OKT3 is muromumab. Absolute lymphocyte count was tested at D3D28.

| | BTI<br>N=17 |
|---|---|
| Delay between 1st vaccine dose and onset of symptoms (days) | 256 (232; 284) |
| Delay between 3rd vaccine dose and onset of symptoms (days) | 87 (65; 117) |
| Delay between onset of symptoms and positive SARS-CoV-2 PCR (days) | 2.5 (1.4; 3.6) |
| Mild disease | 16 (94) |
| Moderate disease / Oxygenorequerance | 1 (6) |
| Severe disease / Intensive care requirement | 0 |
| COVID-19 Death | 0 |
| Cough | 7 (41) |
| Fever | 7 (41) |
| Rhinitis | 6 (35) |
| Diarrhoea | 3 (18) |
| Myalgia | 3 (18) |
| Altered status | 3 (18) |
| Dyspnoea | 2 (12) |
| Vomiting | 2 (12) |
| Odynophagia | 2 (12) |
| Headache | 1 (6) |
| Anosmia | 1 (6) |
| Abdominal pain | 1 (6) |
| Curative monoclonal antibodies | 14 (83) |
| Variant identification performed | 4 (24) |
| Delta variant | 4/4 (100) |

table S2. Characteristics of symptomatic SARS-CoV-2 BTI.

The table includes the 17 cases of symptomatic BTI cases diagnosed after D3D28. Data are geometric means (95% CI) for continuous variables, and n (%) for binary variables.

| | D2D28 | D3D28 | p-value |
|---|---|---|---|
| **RBD IgG** | | | |
| n | 51 | 53 | |
| Concentration (IU/mL) | 8.33 (<5.7;13.3) | 35.1 (16.6; 73.9) | <0.0001 |
| n > 5.4 IU/mL | 21 (41) | 27 (51) | |
| **RBD IgG avidity** | | | |
| n | 16 | 27 | |
| $1/k_{off}$ $(1/s^{-1})$ | 616 (325; 1166) | 3034 (1816; 5070) | 0.0052 |
| **Wuhan $NT_{50}$** | | | |
| n | 51 | 53 | |
| $NT_{50}$ (IU/mL) | <77 (<77; <77) | 102 (<77; 146) | <0.0001 |
| n > 77 IU/mL | 1 (2) | 24 (45) | |
| **S1 IFNγ** | | | |
| n | 46 | 49 | |
| SFC per $10^6$ PBMC | 9.24 (5.58; 15.3) | 9.74 (6.12; 15.5) | 0.65 |
| n > 50 SFC / $10^6$ PBMC | 7 (15) | 9 (18) | |
| **S2 IFNγ** | | | |
| n | 46 | 49 | |
| SFC per $10^6$ PBMC | 12.6 (7.55; 21.0) | 16.1 (9.39; 27.7) | 0.50 |
| n > 50 SFC / $10^6$ PBMC | 11 (24) | 12 (24) | |

#### table S3. Humoral and cellular immune responses at D2D28 and D3D28.

The table includes the 53 patients who remained from free of symptomatic BTI at D3D28. Avidity was measured for sample with RBD IgG > 5.4 IU/mL. Data are geometric means (95% CI) for continuous variables, and n (%) for binary variables. Variables were compared with paired Wilcoxon signed rank test. RBD: receptor binding domain. NT50: 50% neutralizing antibody titer. SFC: spot forming cells. PBMC: peripheral blood mononuclear cells.

| | ALL N=53 | No BTI N=36 | BTI N=17 | BTI versus no BTI Odds ratio (95% CI) | p-value |
|---|---|---|---|---|---|
| **Immune parameters at D3D28** | | | | | |
| RBD IgG (IU/mL) | 35.1 (16.6; 73.9) | 58.5 (22.7; 151) | 11.9 (3.86; 36.6) | (log) 0.57 (0.31; 0.97) | 0.05 |
| RBD IgG avidity ($1/k_{off}$) ($1/s^{-1}$) | 3034 (1816; 5070) | 4103 (2456; 6853) | 1055 (243; 4581) | (log) 0.07 (0.003; 0.61) | 0.04 |
| Wuhan $NT_{50}$ (IU/mL) | 102 (<77; 146) | 136 (83.9; 221) | < 77 (<77; 78.0) | (log) 0.15 (0.02; 0.64) | 0.027 |
| S1 IFNγ (SFC/$10^6$ PBMC) | 9.74 (6.12; 15.5) | 10.4 (5.67; 19.0) | 8.43 (3.99; 17.8) | (log) 0.83 (0.34; 2.00) | 0.7 |
| S2 IFNγ (SFC/$10^6$ PBMC) | 16.1 (9.39; 27.7) | 28.3 (14.4; 55.5) | 5.27 (2.30; 12.1) | (log) 0.30 (0.11; 0.69) | 0.009 |
| **Demographic data** | | | | | |
| Age (years) | 58.6 (54.7; 62.8) | 60.1 (54.8; 66.0) | 55.5 (50.3; 61.3) | 0.96 (0.92; 1.01) | 0.13 |
| Male gender (%) | 32 (60) | 22 (61) | 10/17 (59) | 0.91 (0.28; 3.02) | 0.9 |
| Body mass index (kg/m²) | 25.0 (23.6; 26.5) | 24.6 (22.9; 26.5) | 25.9 (23.3; 28.8) | 0.92 (0.22; 3.43) | >0.9 |
| **Comorbidities** | | | | | |
| Hypertension (%) | 45 (85) | 29 (81) | 16 (94) | 3.86 (0.61; 75.6) | 0.2 |
| Diabetes (%) | 22 (42) | 17 (47) | 5 (29) | 0.47 (0.13; 1.54) | 0.2 |
| Cardiovascular disease (%) | 15 (28) | 12 (33) | 3 (18) | 0.43 (0.09; 1.64) | 0.2 |
| Chronic kidney insufficiency (%) | 3 (6) | 2 (6) | 1 (6) | 1.06 (0.05; 11.9) | >0.9 |
| Cancer (%) | 12 (23) | 8 (22) | 4 (24) | 1.08 (0.25; 4.11) | >0.9 |
| **Transplantation data** | | | | | |
| Transplantation rank > 1 (%) | 5 (9) | 4 (11) | 1 (6) | 0.67 (0.03; 5.69) | 0,7 |
| Time from transplantation to vaccination (years) | 7.1 (5.3; 9.5) | 6.6 (4.6; 9.6) | 8.3 (5.1; 14) | 1.02 (0.94; 1.10) | 0.7 |
| Estimated filtration rate – CKD EPI (mL/min) | 49.9 (45.3; 55.0) | 52.6 (46.7; 59.2) | 44.6 (37.6; 52.9) | 0.97 (0.93; 1.00) | 0.10 |
| OKT3 (%) | 4 (8) | 3 (8) | 1 (6) | 0.71 (0.03; 6.09) | 0.8 |
| Anti-IL2 receptor (%) | 27 (51) | 21 (58) | 6 (35) | 0.86 (0.26; 2.90) | 0.8 |
| Anti-thymocyte (%) | 20 (38) | 11 (31) | 9 (53) | 1.31 (0.37; 4.50) | 0.7 |
| Unknown induction treatment (%) | 2 (4) | 1 (3) | 1 (6) | - | - |
| Triple immunosuppressive therapy (%) | 32 (60) | 20 (56) | 12 (71) | 1.93 (0.58; 7.08) | 0.3 |
| Steroids (%) | 40 (75) | 26 (72) | 14 (82) | 1.79 (0.46; 8.98) | 0.4 |
| Mycophenolate mofetil (%) | 27 (51) | 17 (47) | 10 (59) | 1.60 (0.50; 5.30) | 0.4 |
| Azathioprine (%) | 14 (26) | 10 (28) | 4 (24) | 0.80 (0.19; 2.92) | 0.7 |
| Tacrolimus (%) | 34 (64) | 25 (69) | 9 (53) | 0.50 (0.15; 1.63) | 0.2 |
| Ciclosporine (%) | 8 (15) | 5 (14) | 3 (18) | 1.33 (0.25; 6.22) | 0.7 |
| Everolimus (%) | 15 (28) | 9 (25) | 6 (35) | 1.64 (0.46; 5.71) | 0.4 |
| Absolute lymphocyte count (/µL) | 1214 (1056; 1395) | 1270 (1064; 1515) | 1093 (874; 1367) | (log) 0.2 (0.01; 3.63) | 0.3 |

#### table S4. Univariate analysis assessing the relation between breakthrough infections and clinical or immunological variables

The table includes the 53 patients who remained free of symptomatic BTI at D3D28, including 17 patients who developed symptomatic BTI and 36 patients who did not develop symptomatic BTI after D3D28. Data are geometric means (CI 95%) for continuous variables, and n (%) for binary variables. All demographic and clinical criteria and immunological parameters at D3D28 were included in univariate analysis. Factors associated with the occurrence of BTI was analyzed using univariate logistic regression; odd ratios and p-values are reported. BTI: breakthrough infection. CI: confidence interval. RBD: receptor binding domain. $NT_{50}$: 50% neutralizing antibody titer. SFC: spot forming cells. PBMC: peripheral blood mononuclear cells.

| | RBD IgG | RBD IgG avidity | Wuhan $NT_{50}$ | S1 IFNγ | S2 IFNγ |
|---|---|---|---|---|---|
| **Demographic data** | | | | | |
| Age | **p=0.015; -0.032 (0.013)** | p=0.255 | **p=0.012; -0.016 (0.006)** | p=0.105 | p=0.156 |
| Male gender | p=0.369 | p=0.236 | p=0.573 | p=0.547 | p=0.861 |
| Obesity | p=0.452 | p=0.156 | p=0.090 | p=0.587 | p=0.444 |
| **Comorbidities** | | | | | |
| Hypertension | p=0.070 | p=0.183 | **p=0.011; -0.544 (0.207)** | p=0.106 | p=0.280 |
| Diabetes | p=0.463 | p=0.376 | p=0.851 | p=0.917 | p=0.881 |
| Cardiovascular disease | p=0.143 | p=0.539 | p=0.148 | p=0.176 | p=0.112 |
| Chronic kidney insufficiency | p=0.286 | **p=0.040; -1.167 (0.537)** | p=0.190 | p=0.334 | p=0.353 |
| Cancer | p=0.701 | p=0.917 | p=0.683 | p=0.323 | p=0.172 |
| **Transplantation data** | | | | | |
| Transplantation rank > 1 | p=0.226 | p=0.719 | p=0.351 | p=0.185 | p=0.807 |
| Time from transplantation to vaccination | p=0.197 | p=0.717 | p=0.189 | **p=0.026; 0.046 (0.020)** | **p=0.002; 0.046 (0.014)** |
| Estimated filtration rate – CKD EPI | **p=0.019; 0.021 (0.009)** | p=0.465 | **p=0.024; 0.010 (0.004)** | **p=0.005; 0.025 (0.008)** | p=0.086 |
| Anti-thymocyte | p=0.269 | p=0.691 | p=0.270 | p=0.610 | **p=0.045; 0.487 (0.236)** |
| Anti-IL2 receptor | p=0.168 | p=0.244 | p=0.219 | p=0.998 | p=0.272 |
| OKT3 | p=0.567 | p=0.160 | p=0.758 | p=0.374 | p=0.144 |
| Triple immunosuppressive therapy | p=0.791 | p=0.888 | p=0.904 | p=0.579 | p=0.820 |
| Steroids | p=0.617 | p=0.982 | p=0.518 | p=0.463 | p=0.979 |
| Mycophenolate mofetil | p=0.211 | p=0.231 | p=0.295 | **p=0.031; -0.684 (0.309)** | p=0.073 |
| Azathioprine | p=0.065 | p=0.299 | **p=0.046 ; 0.353 (0.172)** | **p=0.012; 0.890 (0.343)** | **p=0.044; 0.519 (0.251)** |
| Tacrolimus | p=0.208 | p=0.454 | p=0.271 | p=0.993 | p=0.254 |
| Ciclosporine | p=0.255 | p=0.270 | p=0.448 | p=0.420 | p=0.911 |
| Everolimus | p=0.519 | p=0.988 | p=0.399 | p=0.623 | p=0.387 |
| Absolute lymphocyte count | p=0.110 | **p=0.027 ; <0.001 (<0.001)** | p=0.068 | p=0.106 | **p=0.007; 0.001 (<0.001)** |

#### table S5. Linear regression assessing the association between immune parameters and demographical or clinical variables.

The table includes the 53 patients who remained free of symptomatic BTI at D3D28. Linear regression assessing the association between immune parameters and demographical or clinical variables. $p<0.05$ are highlighted in bold. Obesity was defined as BMI > 30 kg/m². Each induction treatment was compared to the other two treatments. Coefficient (Standard error) are reported if $p<0.05$.

| | No BTI n=36 | BTI n=17 | p-value |
|---|---|---|---|
| **B.1.617.2 Delta RBD IgG mean fluorescence intensity** | | | |
| n | 35 | 17 | |
| MFI | 1059 (419; 2680) | 182 (59.3 ; 560) | 0.10 |
| **B.1.617.2 Delta neutralizing antibody titer** | | | |
| n | 36 | 17 | |
| n > 77 IU/mL | 4 (11) | 0 | 0.29 |
| **Omicron BA.1 neutralizing antibody titer** | | | |
| n | 36 | 17 | |
| n > 77 IU/mL | 1 (3) | 0 | >0.99 |
| **Omicron BA.2 neutralizing antibody titer** | | | |
| n | 36 | 17 | |
| n > 77 IU/mL | 4 (11) | 0 | 0.29 |

table S6. Binding and neutralizing antibody titers by variant of concern after three vaccine doses (D3D28).

The table includes the 53 patients who remained free of symptomatic BTI at D3D28, including 17 patients who developed symptomatic BTI and 36 patients who did not develop symptomatic BTI after D3D28. Data are geometric means (95% CI) for continuous variables, and n (%) for binary variables. The lower limit of detection for neutralizing antibody titer IgG was 77 IU/mL. BTI: breakthrough infection. RBD: receptor binding domain. MFI: mean fluorescence intensity. $NT_{50}$: 50% neutralizing antibody titer. Variables were compared with Wilcoxon-Mann-Whitney test.
